# Evaluating adults’ dental caries burden through the DMFT index: results from the Tanzanian 5^th^ national oral health survey

**DOI:** 10.1101/2024.06.28.24309650

**Authors:** Kasusu Nyamuryekung’e, Hawa Mbawalla, Matilda Mlangwa

**Affiliations:** Department of Orthodontics, Pedodontics and Community Dentistry, School of Dentistry, Muhimbili University of Health and Allied Sciences, Dar es Salaam, Tanzania

**Author notes:** Corresponding author (KKN).

## Abstract

**Aim:** Dental caries remains the most prevalent chronic disease worldwide, affecting adults and children across all regions. The DMFT index, representing the number of decayed, missing, and filled teeth, serves as a fundamental metric in oral epidemiology, providing an objective quantification of dental caries prevalence and severity. The aim of the current study is to evaluate the dental caries burden of the adult Tanzanian population through the DMFT index.

**Methods:** A national pathfinder survey was conducted in mainland Tanzania using a cross-sectional design. The survey encompassed fourteen districts across thirteen regions. Site selection followed the World Health Organization’s (WHO) basic oral health survey methods, utilizing a modified stratified-cluster sampling approach. Electronic questionnaires were administered to all adult participants prior to their dental examinations. The questionnaires collected data on participants’ sociodemographic information, tooth brushing practices, and dentition status. Dental examinations were conducted in accordance with WHO standard criteria. Data cleaning and analysis were performed using SPSS version 23. Frequencies were calculated to determine the proportions of participants’ mean and separate DMFT (Decayed, Missing, and Filled Teeth) components. Bivariate associations were examined using Student’s t-test and ANOVA to compare participants’ DMFT components with their sociodemographic characteristics and oral health practices.

**Results:** The study surveyed a total of 1,386 participants aged 30-34, 35-44 and 50+ comprising 713 females (51.4%) with most participants (49.1%) aged 50 years or older. The mean DMFT in the studied population was 4.63 (SD 5.4) with a dental caries experience of 76.6%. The Missing teeth component constituted the largest portion of the mean DMFT (52.4%), followed closely by the Decayed teeth component (45.8%). Tooth fillings were found in just 3.5% of those with dental caries experience. The mean DMFT score was significantly higher among females (5.31 ± 5.79), those aged 50 or more (5.93 ± 6.26) and with lowest level of education (5.78 ± 6.68). Brushing less than once daily (p<0.001), not using toothpaste (p<0.001) and not using a toothbrush daily (p<0.05) were all associated with increased DMFT scores.

**Conclusion:** Dental caries is a significant public health issue in Tanzania. The predominance of missing and decayed teeth highlights the critical gaps in treatment and dental care service accessibility. There is a pressing need for increased public health investment in preventive and restorative dental care and enhanced community education on the importance of maintaining dental health.

## Introduction

Oral health is a critical component of overall well-being, influencing not only the physical ability to perform essential functions such as speaking, chewing, and smiling, but also impacting social interaction and quality of life. It is a significant determinant of overall health and quality of life, particularly among adults, where the burden of oral diseases such as dental caries has far-reaching clinical, economic, and sociodemographic implications. According to the World Health Organization (WHO), regular national oral health surveys are crucial for gathering data that inform the planning and implementation of effective oral health services, and for monitoring trends in oral health across populations(1). These surveys provide essential insights into the prevalence and impact of oral diseases, which are crucial for global and regional health comparisons.

Dental caries and periodontal diseases remain the most prevalent chronic diseases worldwide, affecting adults across all regions, including Africa. Dental caries leads to significant clinical consequences such as pain, discomfort, and functional limitations, impacting dietary choices and nutritional intake (2). The economic implications are equally substantial, with high treatment costs and productivity losses due to dental diseases, straining both individual and public healthcare resources (3). In Sub Saharan Africa, where access to dental care is limited, the economic burden is exacerbated by out-of-pocket expenditures, which are prohibitively high for many households (4,5).

The DMFT index, representing the number of decayed, missing, and filled teeth, serves as a fundamental metric in oral epidemiology. Particularly critical in the context of Low- and Middle-Income Countries (LMICs), the DMFT index provides an objective quantification of dental caries prevalence and severity. In regions where resources for health surveillance are often limited, the DMFT index offers a straightforward, cost-effective means to assess and monitor oral health trends over time. It highlights the gaps in oral health services and the urgent need for interventions to address them.

The DMFT status of African adults reveals significant challenges in oral health, characterized by high levels of untreated dental caries and tooth loss, with very few filled teeth. This trend is largely attributed to limited access to access to preventive and curative dental services, influenced by both geographic and financial barriers (6). Studies show that adults from lower socioeconomic backgrounds are more likely to experience higher DMFT scores, indicating a greater prevalence of untreated caries and tooth loss (7,8). In rural areas, the lack of dental infrastructure exacerbates these issues, resulting in higher DMFT scores compared to urban populations where access to dental services is relatively better (9).

In Tanzania, as in many parts of sub-Saharan Africa, oral health issues are often overlooked in health policy discussions, despite their profound implications on societal well-being and economic stability. Studies within the region have shown that dental caries and other oral health conditions disproportionately affect adults in lower socioeconomic groups, contributing to and exacerbating existing health disparities (8). Thus, assessment of the DMFT status and its sociodemographic correlates is essential not only for understanding current health challenges but also for planning effective preventive and curative strategies tailored to the specific needs of the population. The aim of the current study is to evaluate the dental caries burden of the adult Tanzanian population through the DMFT index, providing valuable insights into the overall impact of dental caries in this setting.

## Materials and methods

### Study design and sites

A national pathfinder survey was conducted in mainland Tanzania, using a cross-sectional design that encompassed fourteen districts. Sites were selected according to the World Health Organization’s (WHO) basic oral health survey methods, employing a modified stratified-cluster sampling approach(1). This methodology ensures the inclusion of diverse population subgroups representing varying levels of oral disease. Specifically, our study involved selecting clusters from cosmopolitan cities, towns, and rural areas. According to the WHO guidelines, a minimum of twelve sites is recommended, split equally among metropolitan areas, large towns, and rural settings. For Tanzania, we expanded this number to fourteen to adequately represent its geographical and demographic diversity.

Tanzania’s 26 regions are distributed across five administrative zones: northern, coastal, central, lake, southern highlands, and western. For our study, three cosmopolitan cities—Dar es Salaam, Arusha, and Mbeya—were purposively chosen based on their size and geographical importance, contributing a total of four cosmopolitan clusters. In addition, regions from each zone were selected based on geographical characteristics and fluoride levels, leading to the inclusion of four regions (Mara, Kigoma, Tanga, and Mtwara) designated as urban clusters. Combining these with the cosmopolitan clusters resulted in eight urban clusters. Additionally, six other regions (Morogoro, Ruvuma, Songwe, Mwanza, Singida, and Tabora) were designated as rural clusters, each contributing one cluster. Thus, the final composition of the study included six rural clusters, four urban clusters, and four cosmopolitan clusters, as detailed in Table 1, which outlines the cluster selection process. Within each district, one ward was randomly selected for inclusion into the study.

**Table 1:**
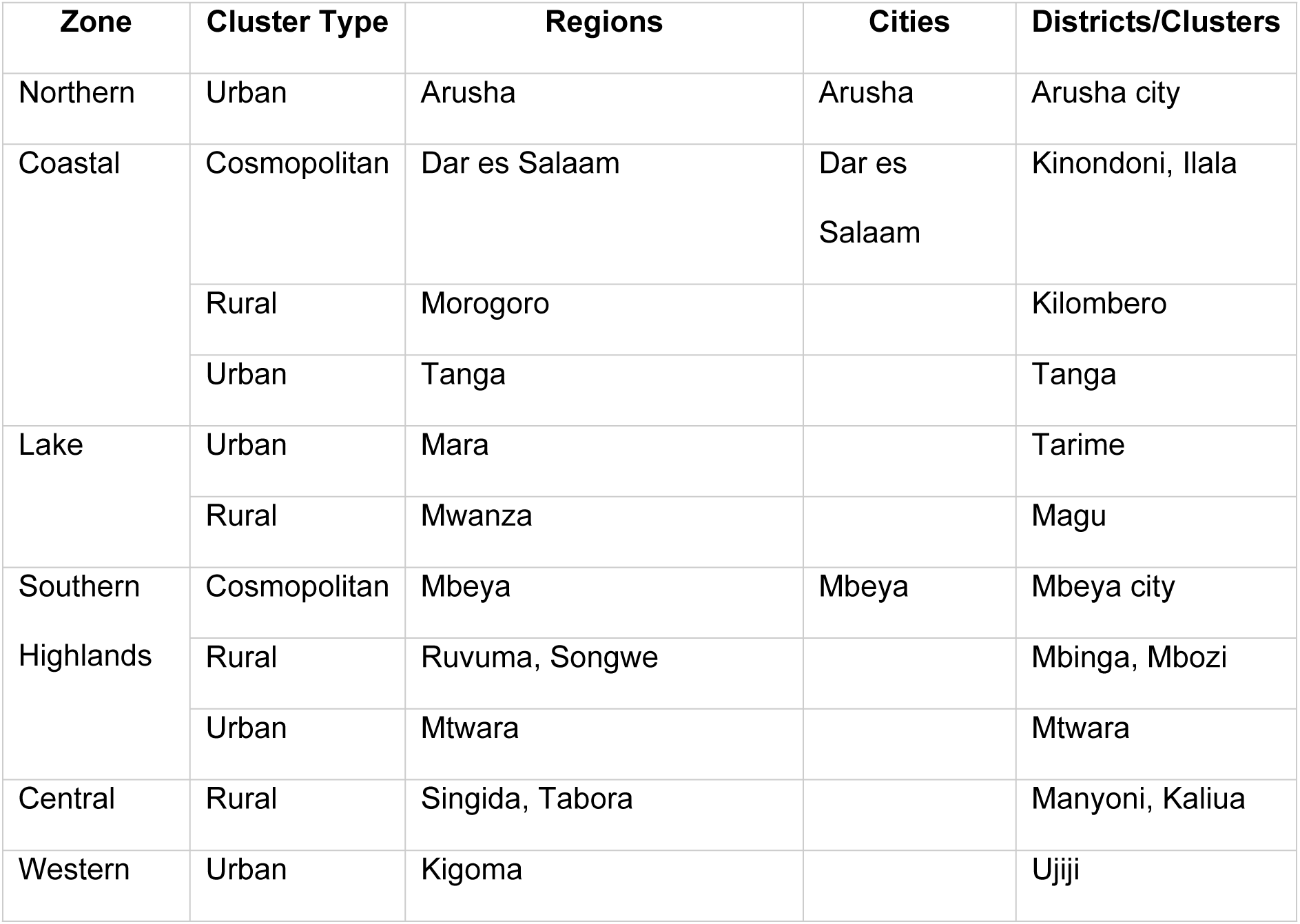
Sampling process for inclusion of participants.

### Sample size and sampling process

We structured the sampling process across six rural and eight urban clusters, stratified by age and sex in alignment with WHO guidelines. Target adult age groups included 30-44, and 50+ years. Following the WHO national pathfinder guidelines, we selected streets randomly within designated wards and recruited 100 adults from each age bracket—50 from the 30–44-year group and 50 from the 50+ group. We continued household visits street by street until achieving the necessary sample size.

### Variables and data collection tools

We measured all variables in accordance with WHO criteria, focusing on a range of indicators including socio-demographics, assessment of dental caries, and oral health practices (1). Data collection was conducted in pairs comprising of a recorder and examiner. Dental caries was assessed using DMFT criteria and was recorded using digitized WHO clinical record forms. Interviewer administered questionnaires were utilized to solicit information on sociodemographic characteristics and oral health practices. Dental caries examination was performed using mouth mirrors under natural lighting conditions. Additional equipment included sterilization devices, storage containers for instruments, gloves, masks, gauze, and refuse bags. During interviews and examinations, participants were seated on standard chairs.

### Reliability and validity

We pre-tested all data collection tools to verify their accuracy, logic, and practicality, including the estimated time required for examining and interviewing participants across different age groups. All investigators and research assistants underwent a comprehensive two-day training at the Muhimbili University of Health and Allied Sciences (MUHAS) Dental School to familiarize themselves with the survey procedures. To enhance validity, examiners were calibrated against senior clinicians and participated in exercises to ensure inter- and intra-examiner consistency after mastering the clinical scoring process.

The WHO (2013) oral health questionnaire, already validated in previous studies including in Tanzania, was translated into Swahili and piloted with 10 adults and 10 children, with only minor modifications made to the wording while preserving the original meaning. Data collectors received detailed information sheets outlining all variables and their operational definitions. To maintain data integrity, all collected information was reviewed daily, and each study team included a research supervisor responsible for monitoring data quality and accuracy.

### Data collection process

We compiled a list of potential villages and streets for study site selection. A district dental officer from each cluster assisted, guided, and facilitated access to these sites. Data collection was carried out by four teams, each consisting of two pairs of examiners and recorders and one supervisor.

All examination instruments were decontaminated using antiseptics, cleaned with liquid soap, and rinsed with water before being sterilized and stored in clinical drums for reuse. We adhered to standard aseptic techniques, which included dusting and disinfecting work surfaces, and the use of gloves and face masks. All used materials such as gauze, gloves, and masks were disposed of in hospital refuse bags and incinerated at nearby health facilities.

For data collection, electronic questionnaires were administered to all adults prior to the dental examination. We utilized REDCAP (Research Electronic Data Capture) software to document both questionnaire responses and clinical data. Research supervisors from each team regularly reviewed and then uploaded the collected data to the REDCAP servers of the Muhimbili University of Health and Allied Sciences (MUHAS). Data was collected for about one month, from 5^th^ February to16^th^ March 2020.

### Data Analysis

Data was transformed to allow computation of sum indices where applicable, including for DMF-T. All participants that had a DMFT score > 0 was categorised as having dental caries experience. Residence was categorized as either urban, rural, or cosmopolitan. Frequencies were generated to determine proportions of participants’ mean and separate DMFT components. Bivariate associations using student’s t-test and ANOVA were used to compare participants’ DMFT components with sociodemographic characteristics and oral health practices.

### Ethical considerations

The study proposal was rigorously reviewed by the WHO and the Borrow Foundation prior to funding approval. Ethical clearance was obtained from the Muhimbili University of Health and Allied Sciences (MUHAS) Institutional Review Board. We also secured the necessary permissions from various authorities, including the Prime Minister’s Office Regional Administration and Local Governments, to access the study sites. Each study team leveraged the official permission letters to facilitate discussions with district authorities for site access.

To ensure participant privacy, both the questionnaire administration and clinical examinations were conducted privately, with precautions taken to prevent overhearing or observation of the procedures by others. We used special coding instead of participant names to maintain confidentiality. Access to data was strictly limited to the study team members. Informed consent was obtained from all participants or their guardians. Verbal consent was obtained from all participants prior to their inclusion in the study. The ten-cell leader or hamlet chairperson escorting the research teams to the respective households witnessed the consenting process. Participants diagnosed with dental diseases or conditions during the study were advised to seek further care from nearby health facilities.

## Results

The study enrolled a total of 1,410 adults, however, due to missing and incomplete data entry, a total of 1,386 participants were included for analysis. Participants had a nearly even distribution of sex, comprising 673 males (48.6%) and 713 females (51.4%). Most of the participants (49.1%) were aged 50 years or older, followed by the age group 35-44 years (30.7%) and 30-34 years (20.2%). A significant portion of the participants resided in rural areas (43.1%), with the remaining split between urban (28.0%) and cosmopolitan (28.9%) settings.

The education levels of the participants varied, with the majority having completed primary education (53.0%), while the remainder had either incomplete primary education (23.7%) or secondary education and above (23.3%). When asked about their perceived teeth status, most participants (69.3%) reported the status of their teeth as good, while the remainder reported the status of their teeth as bad (Table 2).

**Table 2:**
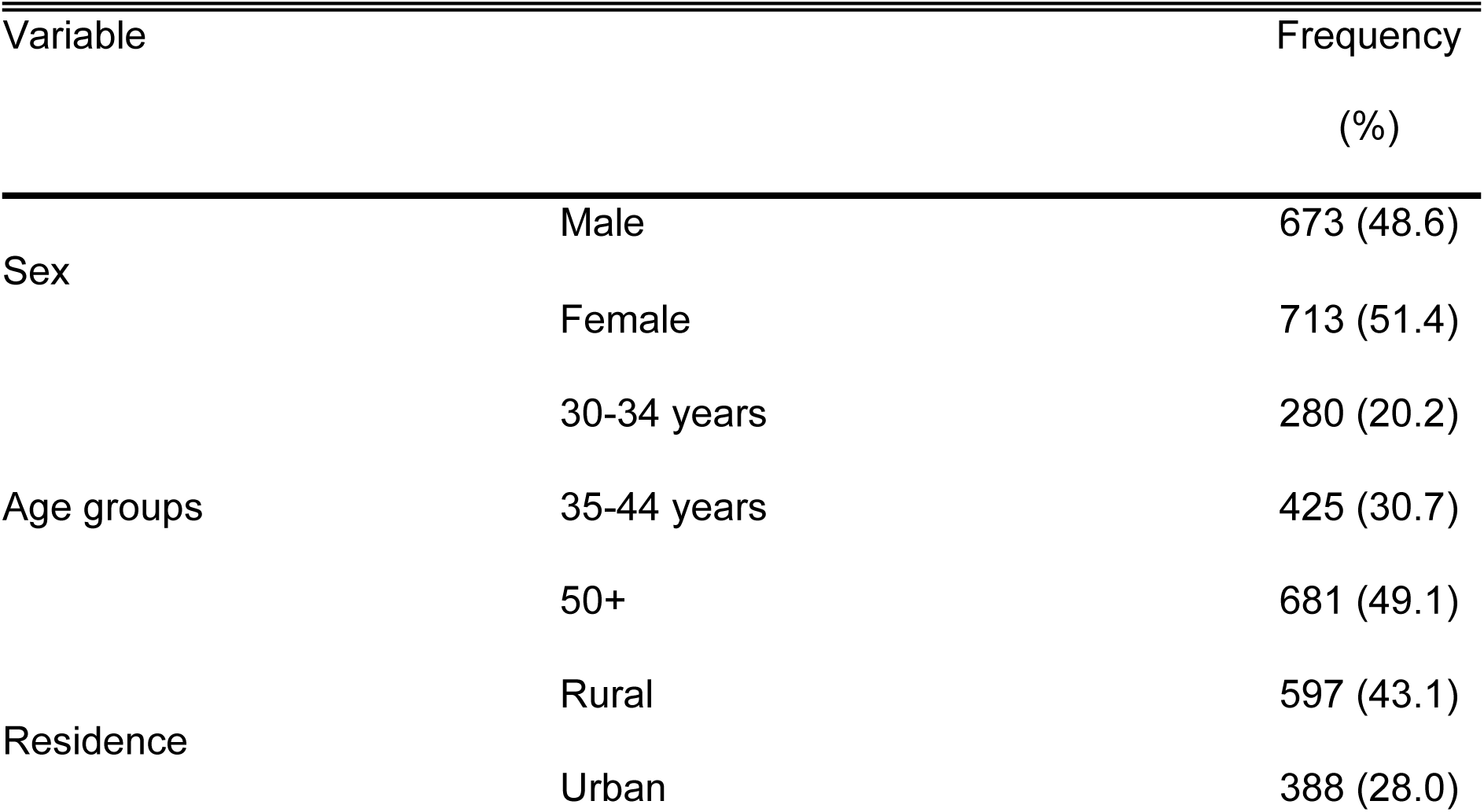
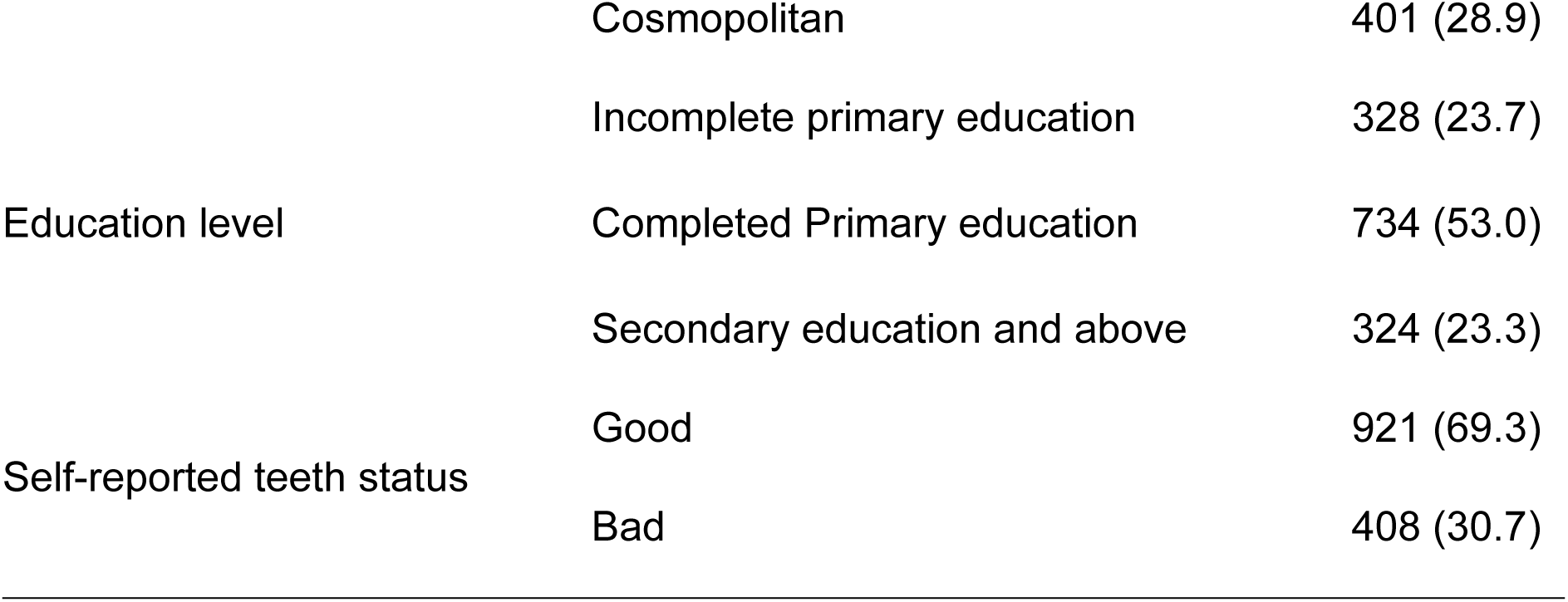
Sociodemographic characteristics of the participants (n = 1,386)

The mean DMFT in the studied population was 4.63 (SD 5.4). The Missing teeth component constituted the largest portion of the mean DMFT (52.4%), followed closely by the Decayed teeth component (45.8%). Tooth fillings accounted for only a small fraction of the overall DMFT, at 4.8%. Overall, more than three-quarters (76.6%) of the population had experienced dental caries (Table 3).

**Table 3:** Participants mean DMFT and dental caries experience.

|  | Frequency (%) | Mean | Std.<br>Deviation |
| --- | --- | --- | --- |
| Decayed teeth | 828 (59.7) | 2.1198 | 2.98880 |
| Filled teeth | 52 (3.5) | .0830 | .52892 |
| Missing teeth | 715 (51.6) | 2.4264 | 4.09781 |
| Dental caries experience (DMFT) | 1,061 (76.6) | 4.6291 | 5.40166 |

The sociodemographic characteristics of the participants were analysed in relation to the individual components of the DMFT index: decayed, missing, and filled teeth. The mean DMFT score was significantly higher among females (5.31 ± 5.79) compared to males (3.91 ± 4.85) (p < 0.001). Females also had higher mean scores for decayed and missing teeth. The mean DMFT score increased with age, showing a significant difference across the age groups (p < 0.001). Participants aged 50 and above had the highest mean DMFT score (5.93 ± 6.26).

The mean DMFT score was similar across different types of residence, although the highest score observed among rural residents (4.86 ± 5.51). The mean DMFT score was significantly higher among participants with incomplete primary education (5.78 ± 6.68) and those reporting to have bad teeth status (7.04 ± 6.60) compared to their counterparts (p < 0.001) (Table 4).

**Table 4:**
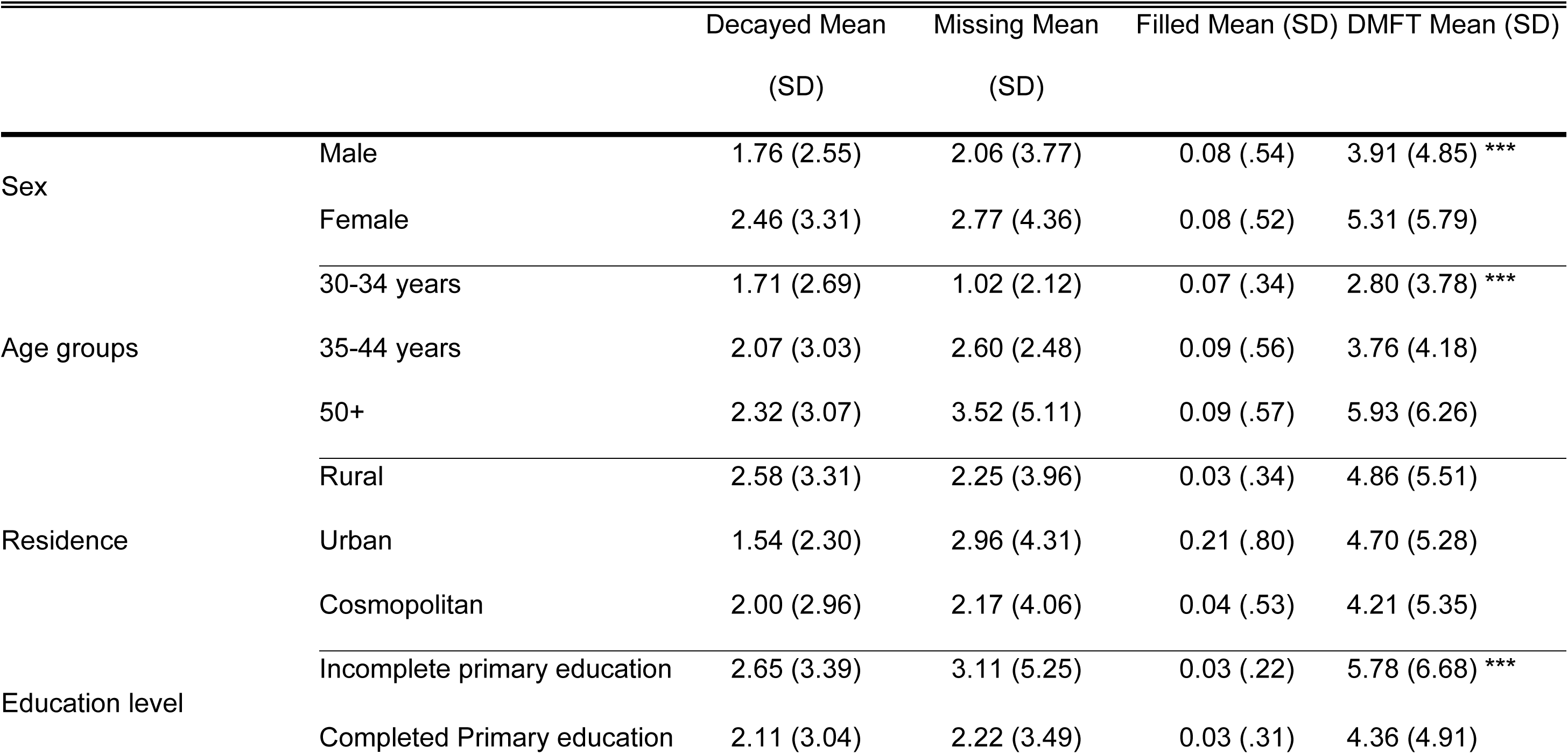
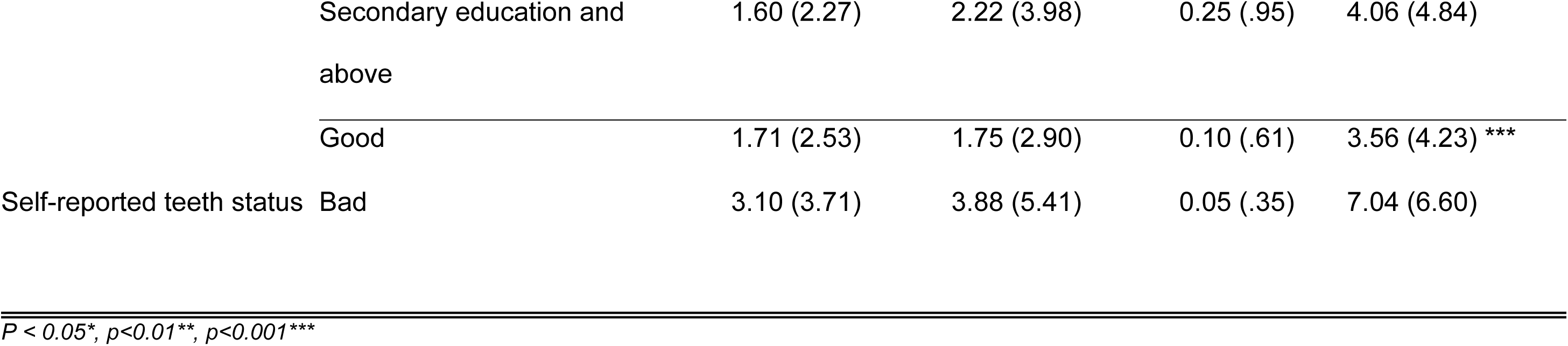
Participants’ sociodemographic characteristics by mean DMFT scores and its components (p-values for independent t-tests and ANOVA).

**Table 5:**
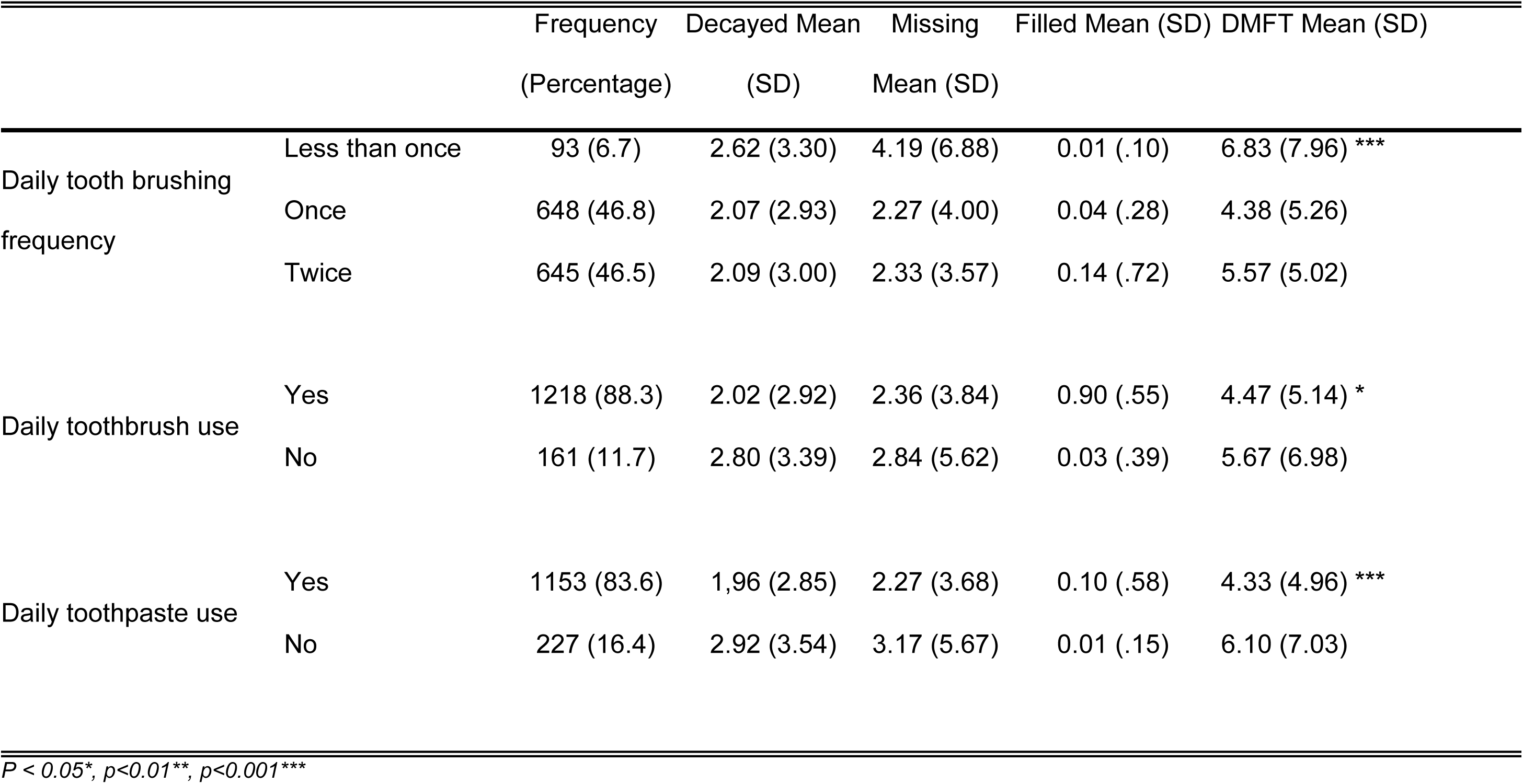
Participants’ tooth brushing practices by mean DMFT scores and its components (p-values for independent t-tests and ANOVA).

The participants’ tooth brushing practices were analyzed in relation to the individual components of the DMFT index: decayed, missing, and filled teeth. The mean DMFT score differed significantly across various tooth brushing frequencies (p < 0.001). Participants who brushed less than once daily had the highest mean DMFT score (6.83 ± 7.96), while those who brushed either once or twice daily had lower mean DMFT scores. Participants who used a toothbrush daily had a significantly lower mean DMFT score (4.47 ± 5.14) compared to those who did not use a toothbrush daily (5.67 ± 6.98) (p < 0.05). Additionally, the mean DMFT score was significantly lower among participants who used toothpaste daily (4.33 ± 4.96) compared to those who did not use toothpaste daily (6.10 ± 7.03) (p < 0.001). (Table 4).

## Discussion

This study provides a comprehensive overview of dental health in the Tanzanian adult population where more than three quarters (76.6%) had dental caries experience. This reflects a global public health challenge and a substantial dental caries burden in this setting. This study reveals a mean DMFT (Decayed, Missing, Filled Teeth) score of 4.63, suggesting a moderate burden of dental diseases. The distribution within the DMFT components was disproportionately skewed towards missing and decayed teeth. Significant associations were observed between sociodemographic characteristics and the DMFT (Decayed, Missing, Filled Teeth) index, illustrating disparities in dental health across different population segments.

The observed prevalence of dental caries experience in this study, at 76.6%, is particularly high even though in alignment with the global burden of dental diseases, which remain a major public health challenge. Globally, there has been a decline in dental caries prevalence in some developed countries due to enhanced preventive measures with more modest improvements in low-resource settings (10).

Nevertheless, current findings remain considerably high even when compared with other studies from low- and middle-income countries (LMICs). A systematic review and meta-analysis revealed the overall prevalence of dental caries in East Africa as 45.7%, with Eritrea having the highest prevalence at 65.2% and Tanzania the lowest at 30.7%. The mean DMFT score for the region is 1.941, with Sudan showing the highest mean DMFT score at 3.146 and Tanzania the lowest at 0.945 (11) This difference may indicate varying degrees of public health intervention effectiveness, differences in dietary habits, fluoride exposure, and access to dental care services across different contexts. Furthermore, the observed discrepancy of current findings compared to the systematic review are to be expected, due to aggregation of data from different population groups and settings. The high prevalence of adult dental caries in our study necessitates integrated preventive and treatment strategies in dental care, particularly in targeting at-risk populations through community-based programs and improving the overall oral health infrastructure. These interventions are critical in reducing the burden of dental caries and should be a priority for health policymakers.

The missing teeth component was the most significant component of the DMFT index at 52.4%, considerably higher than that found in other low- and middle-income countries (LMICs), where the range typically varies more broadly. Studies from other regions in Sub-Saharan Africa and Southeast Asia have reported missing teeth components ranging from 40% to 55%, suggesting a common trend of high tooth loss in these region (12,13). Our findings align with trends observed in the Global Burden of Disease (GBD) study, which highlights tooth loss as a persistent challenge in regions with healthcare disparities. The significant proportion of missing teeth in the studied population indicates a pressing need for enhanced dental health strategies that focus not only on treatment but also on the prevention of initial decay and the promotion of restorative care over extractions.

Decayed teeth constituted a lower but significant portion of the DMFT index at 45.8%, notably higher than global averages (2). This proportion is also higher compared to similar studies conducted in sub-Saharan Africa (14). Our findings suggest a considerable gap in effective dental treatment and possible deficiencies in adequate fluoride exposure, which has been proven to substantially reduce decay rates (15). The high prevalence of untreated decay in this study indicates barriers to accessing dental health services, affordability issues, or a lack of public awareness regarding the importance of regular dental care. There is an urgent need for establishment of enhanced dental health strategies encompassing comprehensive dental care and preventative measures in this setting.

Contrastingly, the filled component of the DMFT was notably low at only 4.8%, substantially lower than what has been reported in other low- and middle-income countries (LMICs). Research by Frencken et al. (2017) in LMICs suggests a slightly higher prevalence of filled teeth, often averaging around 10% to 15% (16). However, our findings are similar to other studies from parts of Africa where access to dental care is extremely limited, with reported filled components as low as 2-3% (4), which highlights the challenges faced in the most resource-constrained settings. The disparity in the filled component of the DMFT observed in this study highlights the urgent need for enhanced dental services and preventive care in the studied population. The significantly low rate of filled teeth not only reflects the lack of access to restorative dental care but also potentially signifies cultural and socioeconomic barriers that prevent Tanzanian populations from seeking treatment. Thus, improving access to dental health services and promoting oral health education remain crucial for increasing the rate of tooth filling utilization in Tanzania and other similar settings.

This study reveals significant associations between sociodemographic characteristics and the DMFT (Decayed, Missing, Filled Teeth) index, illustrating disparities in dental health across different population segments. The data indicate that females had a higher mean DMFT score compared to males, and this difference was statistically significant. This gender disparity is consistent with other studies which suggest that biological, behavioural, and cultural factors may contribute to higher caries prevalence and tooth loss among females (17,18). Moreover, the progression of DMFT scores with age was expected and aligns with global trends, where older age groups typically exhibit more pronounced dental issues due to cumulative exposure to caries risk factors and the natural aging process affecting oral health (19). Participants aged 50 and above displayed the highest mean DMFT scores, underscoring the need for age-specific dental health interventions and preventive measures. Interestingly, the analysis revealed no significant variation in DMFT scores based on types of residence, though rural residents tended to have slightly higher scores. This could suggest underlying factors like differential access to dental care services, dietary habits, and fluoride exposure, which are often less favourable in rural settings (20).

Educational level appeared to play a critical role in dental health outcomes. Participants with incomplete primary education exhibited significantly higher DMFT scores. Education is a well-documented determinant of health, influencing both health behaviours and access to healthcare services, including preventive dental care (21). Lower educational attainment is often associated with inadequate oral hygiene practices, lower health literacy, and reduced utilization of dental care services, which could explain the higher DMFT scores observed in this subgroup. Furthermore, self-reported bad teeth status was strongly associated with higher DMFT scores. This correlation not only reflects subjective oral health assessments but also emphasizes the impact of personal oral health perceptions on overall dental status. Individuals perceiving their dental health as poor might be those experiencing more severe dental problems, indicative of both the extent and progression of untreated dental conditions (22)

The analysis of toothbrushing practices and their association with the DMFT (Decayed, Missing, Filled Teeth) index in this study emphasizes the significant impact of daily oral care routines on dental health outcomes. Participants who engaged in more frequent tooth brushing exhibited substantially lower DMFT scores, demonstrating the effectiveness of regular dental plaque removal in preventing dental caries and subsequent tooth loss. This finding is consistent with previous research that highlights the critical role of tooth brushing frequency as a key preventive measure against tooth decay. Studies have shown that tooth brushing performed at least twice daily significantly reduces caries risk, emphasizing the need for public health initiatives to promote regular brushing habits to maintain dental health (23). Furthermore, the study found that daily use of a toothbrush and toothpaste was associated with lower DMFT scores, with participants using fluoride toothpaste daily exhibiting a notable decrease in their mean DMFT score. This supports the well-documented benefits of fluoride toothpaste in enhancing tooth enamel resistance to acid attacks from bacterial metabolism, reinforcing fluoride’s role as a vital preventive agent against dental caries widely recognized in public health dentistry (24).

The presented findings need to be interpreted while acknowledging some of its limitations. The cross-sectional design of this survey-based study inherently limits its ability to establish causality between oral hygiene practices and DMFT scores, capturing only a snapshot in time and restricting insights into the progression of dental health issues over time. Additionally, the majority of participants being aged 50 years or older (49.1%) might skew results toward dental issues more prevalent in older adults, potentially limiting the applicability of findings to younger populations. Furthermore, the reliance on self-reported measures for oral hygiene practices introduces the risk of bias due to misreporting or memory recall issues, which could affect the accuracy of the correlations between reported hygiene practices and actual oral health outcomes. Lastly, while the study controls for several demographic factors, it may not adequately address other confounding variables such as dietary habits, water fluoridation, and access to dental care, which could significantly influence DMFT scores and overall oral health, thus potentially biasing the study’s results.

Nevertheless, this study employed the WHO pathfinder method to enrol a nationally representative sample, thereby enhancing the generalizability of the findings to the broader population and allowing for an accurate reflection of the national status of oral health, which can effectively inform policy decisions. The use of validated clinical assessment tools and questionnaires ensured the reliability and validity of the data collected, minimizing bias and enhancing the accuracy of dental caries and other dental conditions diagnoses. With a nearly even gender distribution among participants, the study effectively reflected gender differences in oral health, crucial for analysing sex-specific outcomes and tailoring health interventions. Additionally, the inclusion of participants across a wide age range and from diverse geographic settings—rural, urban, and cosmopolitan—added depth to the analysis of age-related and environmental factors affecting oral health, supporting a detailed examination of the impacts of demographic factors on dental health.

## Conclusions

Dental caries is a significant public health issue in Tanzania. The predominance of decayed teeth over filled teeth within the caries-experienced group further highlights the critical gaps in treatment and the need for increased public health investment in dental care and community education about the importance of maintaining dental health. There is a clear need for an integrated approach that combines preventive measures such as increased availability of affordable fluoridated toothpaste and oral health education with improved accessibility to dental services to address this challenge.

## Data Availability

All relevant data are within the manuscript and its Supporting Information files.

## Acknowledgements

We extend our sincere gratitude to the Ministry of Health Tanzania, the President’s Office Regional Administration and Local Government, and the Tanzania Dental Association for their invaluable support and facilitation. Special thanks to the WHO for reviewing the survey proposal, and to the Regional and District Dental and Medical Officers who assisted with logistics and data collection. We are also immensely grateful to all the participants whose cooperation made this study possible. Sincere appreciation to Dr. Joyce Rose Masalu for her tireless dedication and unwavering support towards the conduct of the 5^th^ Tanzanian National Oral Health Survey.

